# Innovative AI models for clinical decision-making: predicting blastocyst formation and quality from time-lapse embryo images up to embryonic day 3

**DOI:** 10.1101/2025.02.15.25322247

**Authors:** A Yanai, A Horie, A Sakurai, S Imakita, M Nakamura, A Ikeda, S Shitanaka, T Ohara, B Nakakita, A Ueda, Y Kitawaki, Y Sagae, A Okunomiya, M Mandai

**Author notes:** **Corresponding author:** Akihito Horie, MD, PhD, Department of Gynecology and Obstetrics, Kyoto University Graduate School of Medicine, 54 Shogoin Kawahara-cho, Sakyo, Kyoto 606-8507, Japan.

## Abstract

Accurate embryo assessment on embryonic day 3 of assisted reproductive technology (ART) is crucial for deciding whether to continue the culture until day 5 (blastocyst stage) or opt for earlier transfer or cryopreservation. Prolonged culture often improves pregnancy outcomes in patients with multiple high-quality embryos, but may offer limited benefits for older patients or those with few available embryos. To address this clinical challenge, analyzing embryo quality in early stages by artificial intelligence (AI) can be useful. We retrospectively analyzed images of 7,111 two-pronuclear embryos (Veeck grade ≤3) using four different time-lapse incubators.

We fine-tuned ImageNet-1k-pretrained NASNet-A Large to automatically classify each time-lapse image into 17 morphological categories, including cell stages and Veeck grades 1–3. This model achieved 95% cell-stage accuracy on the test set. We combined these annotations with age at egg retrieval in a gradient boosting framework (XGBoost) to predict blastocyst formation, good blastocysts, and poor blastocyst + arrested embryos (PBAE). The ROC AUCs were 0.87, 0.88, and 0.87 for blastocyst formation, good blastocysts, and PBAE, respectively, indicating good predictive performance for day 3 embryo assessment. Notably, the PBAE model reached a precision-recall AUC of 0.90, accurately identifying embryos unlikely to benefit from extended culture.

This revolutionary AI prediction model could ensure transparency and addresses the “black box” limitation often associated with AI. By integrating a high-accuracy auto-annotation pipeline with interpretable AI (via SHapley Additive exPlanations), our device-independent approach supports appropriate embryo-specific decisions, potentially reducing unnecessary culture, optimizing workflows, and improving clinical outcomes in ART.

**Capsule:** Artificial intelligence models accurately predicted blastocyst formation and quality using time-lapse images and age on embryonic day 3, supporting clinical decision-making regarding blastocyst culture or early embryo transfer.

**Highlights:** Time-lapse (24–64 hpi) enables accurate day 3-based blastocyst prediction (AUC=0.87). Poor blastocyst + arrested embryo model offers PR AUC=0.90 for early decisions.

Robust across four incubator types and age groups, demonstrating broad utility. SHAP analysis clarifies how morphological features drive AI predictions.

Embryo-specific decisions reduce unnecessary culture and improve patient outcomes.

## Introduction

Assisted reproductive technology (ART) plays an important role in infertility treatment. In 2021, 9% of all births in Japan resulted from ART [1]. Within ART, there are two main timings for embryo transfer: cleavage-stage transfer (on the embryonic day 2 or 3) and blastocyst-stage transfer (on the embryonic day 5 or 6). In patients with a good prognosis, blastocyst**-**stage transfer is usually preferred owing to higher pregnancy and birth rates per transfer [2,3]. According to the Japanese National Registry of ART [1,4], 73% of transfer cycles in 2021 were blastocyst-stage transfers, and the pregnancy rate per embryo transfer was double that of cleavage-stage transfers (41% vs. 20%, respectively). Conversely, in patients with poor prognoses and few available embryos (e.g., over 40 years of age), prolonged culture sometimes fails to produce transferable blastocysts. Several reports [5–7] showed that extending culture to the blastocyst stage did not improve implantation or cumulative pregnancy rates in certain scenarios. Additionally, one report [8] pointed out that the live birth rate of cleavage-stage embryo transfer was double that of blastocyst culture in patients with only one available embryo on embryonic day 3. Thus, cleavage-stage embryo transfer remains an important option for patients with a poor prognosis, such as older patients. In Japan, where donor eggs are difficult to obtain, 47% of patients aged >43 years undergo cleavage-stage transfer [1].

In previous studies [2], the choice between early embryo freezing/transfer and blastocyst transfer was made on a patient-by-patient basis rather than an embryo-by-embryo basis. Embryo-by-embryo optimal selection requires accurate prediction of morphological quality on embryonic day 5 or 6 by embryonic day 3. Although one report [9] showed that static morphological assessment on embryonic day 3 could predict blastocyst formation, the receiver operating characteristic area under the curve (ROC AUC) of 0.78 was considered insufficient for reliable clinical decision-making.

Since 2008, time-lapse embryo culture [10] has enabled the evaluation of morphokinetic parameters, leading to multiple studies reporting predictions of blastocyst formation [11,12,21,13–20] and good blastocyst formation [18,20,22–24] using these parameters. While day 4 predictions of blastocyst formation achieved validated AUCs as high as 0.85 [19], clinically useful predictions at day 3 remained lower, with AUCs of 0.73–0.81 [17,21].

Recent advances have included artificial intelligence (AI) analysis of time-lapse images [25–29], with one reported AI-based model achieving an AUC of 0.82 for predicting blastocyst formation on embryonic day 3 [29]. However, previously reported predictive models [9,17,21,29] face several limitations. First, the prediction accuracy was insufficient for reliable clinical decisions regarding whether extended blastocyst culture or early embryo transfer/freezing on embryonic day 3. Next, the time-lapse parameters required laborious manual annotation and exhibited interrater variability [30]. Last, AI prediction models often lacked interpretability (“black box” issue) and were highly device-dependent, as they were trained using images from only one type of time-lapse incubator [25–29].

This study aimed to develop an accurate AI blastocyst prediction model on embryonic day 3 with high versatility and interpretability for clinical use. To achieve this objective, we analyzed time-lapse images acquired from multiple time-lapse incubators at multiple facilities, constructed models from human-interpretable features, and visualized the AI decision basis using SHapley Additive exPlanations (SHAP) [31,32].

## Materials and Methods

### Overview of the Blastocyst Prediction using AI

An overview is shown in Figure 1. We created a dataset for the blastocyst prediction model (Figure 1A). We fine-tuned the ImageNet-1k pre-trained image analysis AI (NASNet-A Large [33]), using a 17-class embryo image classification dataset to build an AI auto-annotator (Figure 1B). AI extracted features from time-lapse images, and the age at oocyte retrieval was analyzed using a gradient boosting method (XGBoost [34]) to build each prediction model for blastocyst formation, good blastocyst formation, and poor blastocyst + arrested embryos (Figure 1E). Our AI blastocyst prediction model predicted embryonic morphology on embryonic day 5 or 6 based on embryo culture time-lapse images and age at oocyte retrieval.

**Figure 1.**
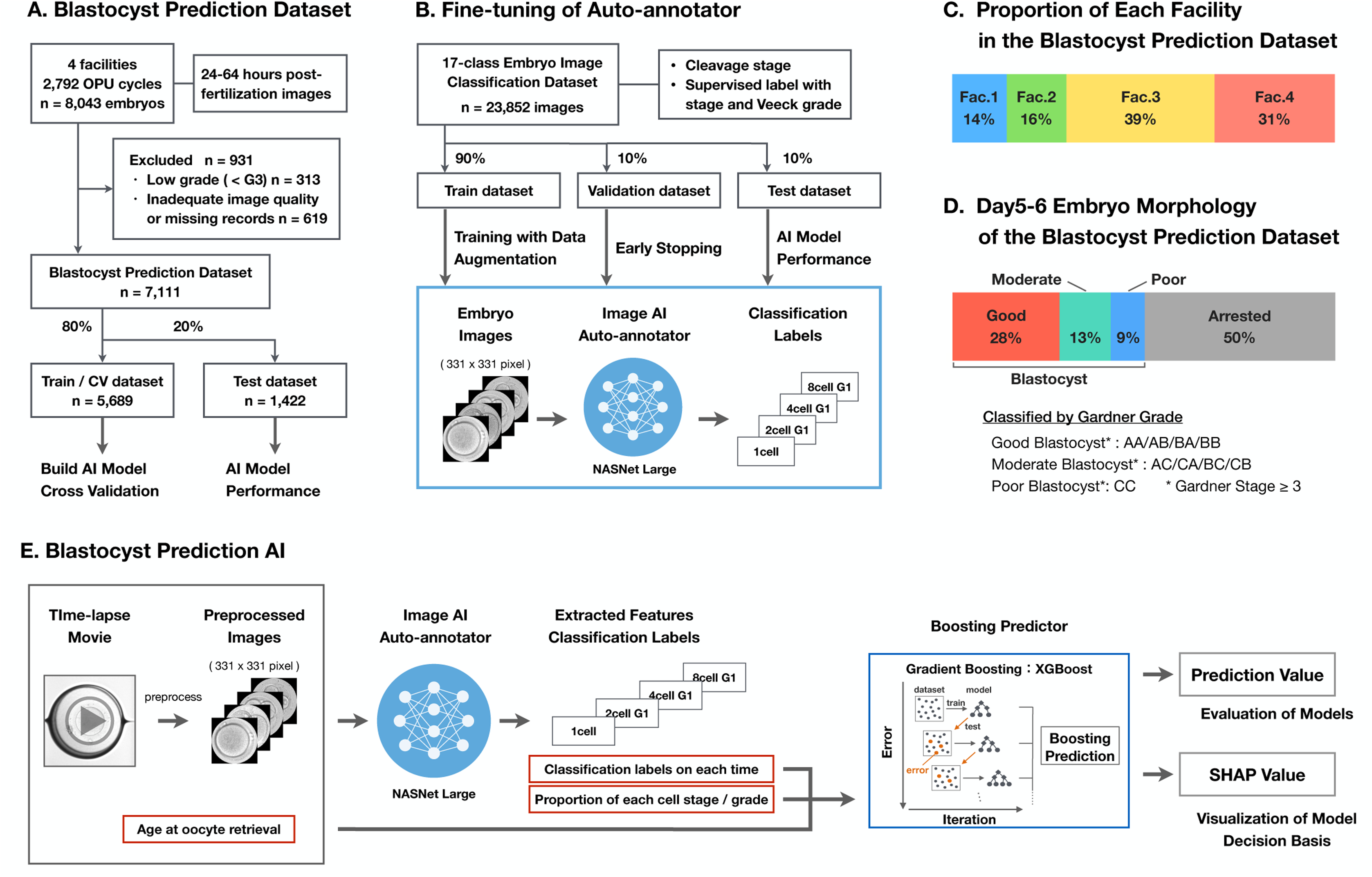
Overview of our blastocyst prediction AI. (**A**) Flowchart of the blastocyst prediction dataset. (**B**) Flowchart of the 17-class embryo image classification dataset and fine-tuning of AI auto-annotator. (**C**) Proportion of each facility in the blastocyst prediction dataset. (**D**) Proportion of embryo morphology on embryonic day 5 or 6 in the blastocyst prediction dataset. (**E**) Schema of the blastocyst prediction AI structure. AI = artificial intelligence, CV = cross-validation, n = number of embryos, Fac. = facility, SHAP = SHapley Additive exPlanations

### Fine-Tuning the AI Auto-Annotator

A 17-class embryo image classification dataset (Figure 2A) was created from 23,852 embryo images with 17-class supervised labels, including 1-cell stage with or without pronuclei (PN) and 2-, 3-, 4-, 5–7-, and 8-cell stages with Veeck grades 1/2/3 [35]. The ImageNet-1k pre-trained image AI, NASNet-A Large [33], was fine-tuned on this dataset to create an AI auto-annotator (Supplementary Text 1). Time-lapse images were analyzed using the AI auto-annotator and classified into 17-class morphological labels. The proportions of each cell stage and Veeck grade were calculated in each embryo as features. The Veeck grade was replaced by the mode frequency (or a better grade if the mode frequency was the same) at each cell stage for each embryo.

**Figure 2.**
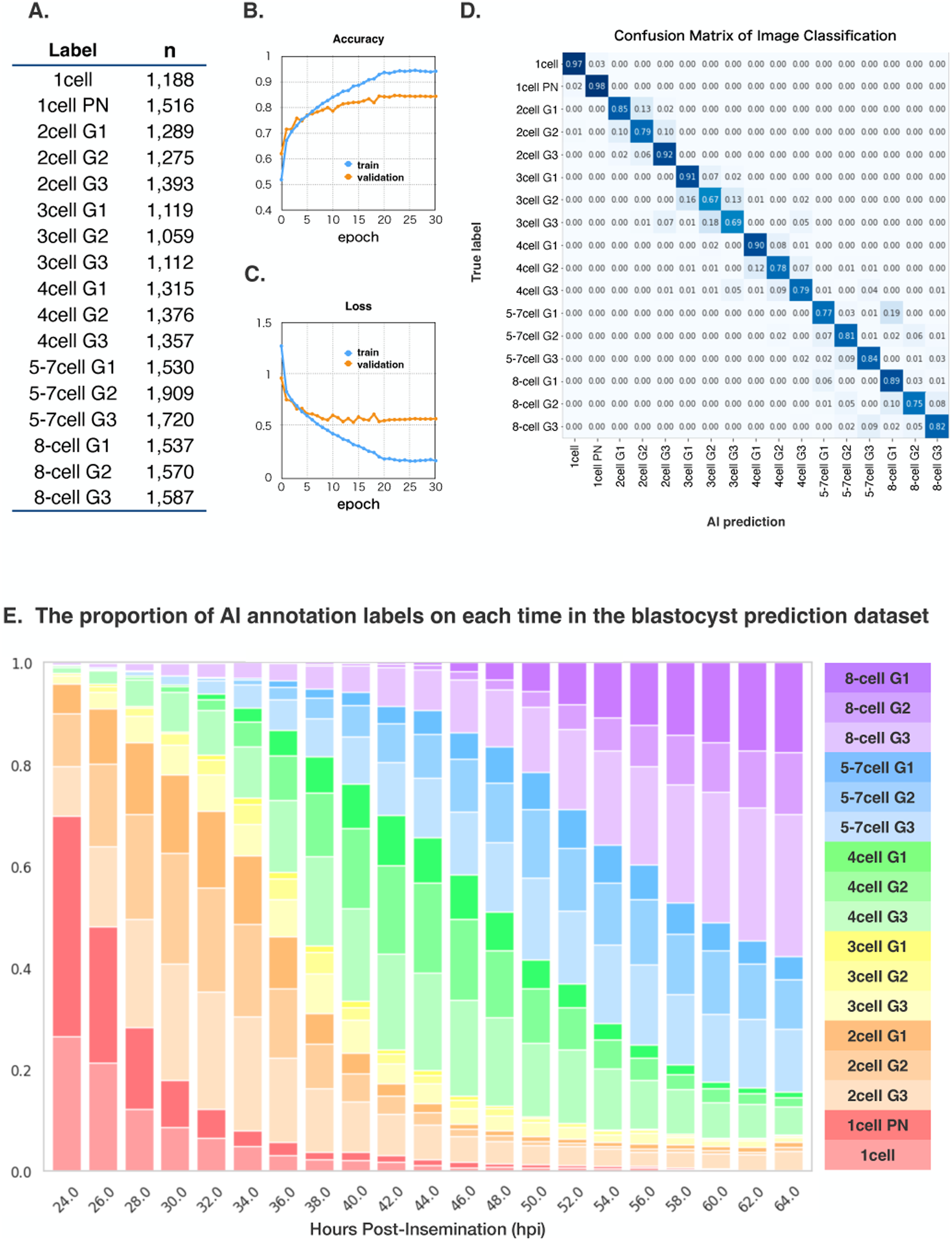
Fine-tuning of the AI auto-annotator. (**A**) Labels and number of images in the 17-class embryo image classification dataset. (**B, C**) Accuracy and loss of fine-tuning. (**D**) Confusion matrix of fine-tuning (17-class image classification). (**E**) Proportion of AI annotation labels at each time point in the blastocyst prediction dataset. AI = artificial intelligence.

### Data Collection and Preparation for the Blastocyst Prediction Dataset

Embryo culture time-lapse images and clinical data recorded in daily practice were retrospectively collected from Kyoto University Hospital and its cooperating facilities from May 2018 to December 2022 (Supplementary Text 1 and Figure 1C).

This study included 2PN embryos with cleavage that were time-lapse cultured, with at least embryonic day 5, single embryo per well. Degenerated and poor embryos with Veeck grades 4 and 5 were excluded. Time-lapse images were collected every 15 min at 24–64 hours post-insemination (hpi). Time-lapse images were collected up to 64 hpi because most day 3 embryo transfer/freezing were performed after 64 hpi. Time-lapse videos were preprocessed and converted to JPG images of 331 × 331 pixels. Embryos with more than 5% image loss were excluded due to missing data, debris, misplacement or significant preprocessing errors. (Supplementary Text 1).

The blastocyst prediction dataset was created by combining the time-lapse images at each time point with the morphological evaluation on embryonic day 5 or 6, age at oocyte retrieval, and facility.

Overall, 8,043 fertilized embryos cultured in four types of time-lapse incubators in 2,792 oocyte retrieval cycles at four facilities were included. After exclusions, 7,111 embryos remained. The dataset was randomly split into 80% for training and cross-validation and 20% (1,422 embryos) for testing (Figure 1A).

Blastocysts were evaluated for Gardner stage and grade; blastocyst formation was defined as Gardner stage ≥3, and arrested embryos were defined as others. Good blastocysts were defined as Gardner grades AA, AB, BA, and BB; poor blastocysts as CC; and others as moderate blastocysts. The morphological evaluation is shown in Figure 1D.

### Building the AI Blastocyst Prediction Models

The AI auto-annotator was used to classify the time-lapse images at each time point into morphological labels, and the proportions of cell stages and Veeck grades were calculated. Time-series labels, proportions, and ages at oocyte retrieval were analyzed using XGBoost and models were built from the training and cross-validation dataset (Supplementary Text 1). The targets for prediction were blastocyst formation, good blastocyst formation, and poor blastocyst + arrested embryos. The models were five-fold cross-validated and tested on a test dataset. The endpoint was ROC AUC and the AUC for precision-recall (PR). For clinical decision-making, a target PR AUC of poor blastocyst + arrested embryos prediction was set at 0.90. Subgroup analyses were performed by facility and age group (<35, 35–39, and ≥40 years).

### Blastocyst Prediction Models from Single or Few Embryos Images

To examine the superiority of time-lapse image analysis over static image analysis, prediction models were created from a single image at 24, 48, or 64 hpi, or three images at 24, 48, and 64 hpi, assuming a once-a-day observation, and ROC AUC values were compared.

In the first type of model, the AI auto-annotator classified static images into morphological annotation labels, and XGBoost built prediction models from single or three labels and age. For the next type of model, the fine-tuned NASNet-A Large was trained on single images (grayscale, 331 × 331 pixels) at 24, 48, and 64 hpi in the training and cross-validation dataset to create prediction models. Additionally, because NASNet-A Large is a color image analysis AI that allows up to three channels of input for each image, we created merged images using 24-, 48-, and 64-hpi images for each channel and trained NASNet-A Large on the merged images to create the prediction models.

### SHAP-Based Model Interpretation

In our prediction models, the AI auto-annotator classified embryo images into human-recognizable morphological labels, and all features inputted to XGBoost were interpretable. However, because XGBoost has dozens to hundreds of decision trees, it is difficult to directly interpret the basis for the prediction models. Therefore, we used SHAP [31,32] to analyze the contribution and predictive direction of each feature in the test dataset. Because SHAP values are positive and negative for positive and negative predictions, respectively, we calculated the absolute mean of SHAP values for each feature and extracted the highly contributing features. The correlation between the value and SHAP value of each contributing feature was visualized and used to interpret the models.

### Statistical Analyses

The model performance was evaluated using ROC and PR analyses and AUCs. The analyses were performed using numpy 1.21.6, scipy 1.7.1, and sklearn 1.0.2 on Python 3.7.15. In the comparison of blastocyst prediction models from time-lapse images and those from single or few embryo images, ROC AUCs were compared, and p-values were calculated using the DeLong test. The DeLong test was performed using the fast DeLong method in Python (https://github.com/yandexdataschool/roc_comparison) [36,37]. The significance level was set at p<0.05; however, because of nine-fold tests, the significance level was set at p<0.0055 with Bonferroni correction.

### Ethical Considerations

This study was conducted in accordance with the principles embodied in the Declaration of Helsinki and was approved by the Ethics Committee of Kyoto University Graduate School and Faculty of Medicine (approval number R3320). Informed consent was obtained in the form of opt-out through the websites of facilities.

## Results

### AI Auto-Annotator Performance

We fine-tuned ImageNet-1k pre-trained image AI NASNet-A Large with the 17-class embryo image classification dataset (Figures 1B and 2A). The training was early stopped at 30 epochs (Figure 2B and C), and the test accuracy was 85%. The confusion matrix (Figure 2D) showed that most misclassifications were due to errors in the Veeck grade, the cell stage accuracy was excellent at 95% (Supplementary Figure 1).

The distribution of AI annotation labels at each time point in the blastocyst prediction dataset showed the progression of cleavage from two to eight cells (Figure 2E). At 24 hpi, the percentage of annotations misclassified as four or more cells owing to fragmentation or other factors was small (2.0%).

Major time-lapse parameters were calculated from the AI annotation labels and compared with those in previous reports [12,13,17,19,38] (Supplementary Table 1). AI-calculated t2 (time to the 2-cell stage) was significantly smaller in the blastocyst formation group than in the arrested embryo group (median: 24.8 vs. 27.0 hpi, p<0.001) and was within the mean or median values in previous reports.

### Blastocyst Formation Prediction Model

Using XGBoost and recursive feature elimination, 88 features were selected for the blastocyst formation prediction model (Supplementary Table 2). The model achieved a test AUC of 0.87 and a five-fold cross-validation mean AUC of 0.86 (Figure 3A). With a Youden’s index [39] threshold of 0.49, the test accuracy reached 0.79. These results demonstrated the model’s robustness in cross-validation and testing.

**Figure 3.**
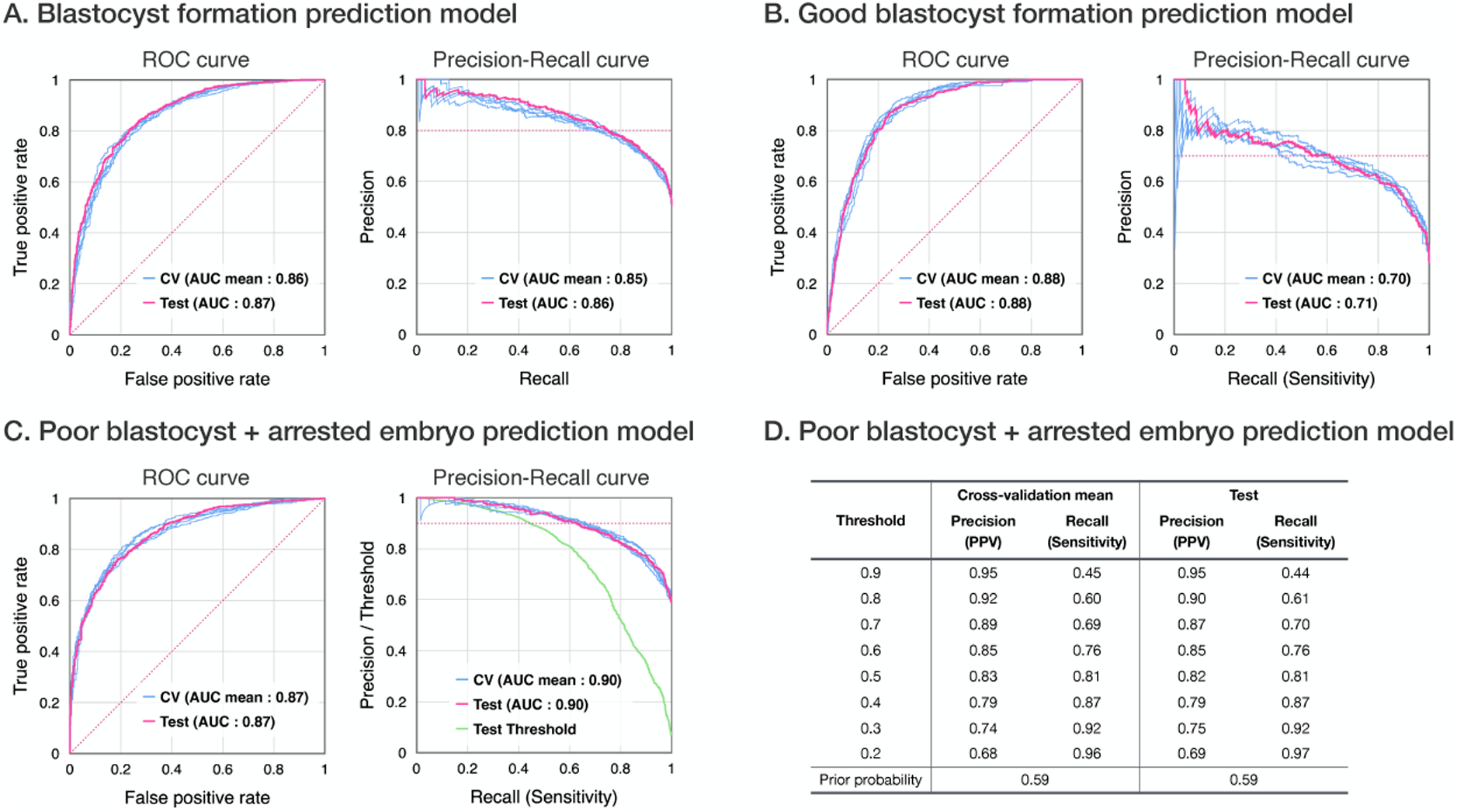
Performance of the prediction models. (**A, B, C**) ROC and precision-recall analyses of three prediction models. The blue lines indicate the results of five-fold validations, whereas the red line denotes the test result. (**D**) Precisions and recalls at each threshold in the poor blastocyst + arrested embryo prediction model. AUC = area under the curve, CV = cross-validation, ROC = receiver operating characteristic, PPV = positive predictive value

### Good Blastocyst Formation Prediction Model

The good blastocyst formation prediction model achieved a test ROC AUC of 0.88 using 156 features. Owing to the low proportion of good blastocysts (28%), the test PR AUC was relatively lower than other models (Figure 3B).

### Poor Blastocyst + Arrested Embryo Prediction Model

The poor blastocyst + arrested embryo prediction model (Figure 3C), using 76 features, achieved a test AUC of 0.87. Its PR AUC reached 0.90, indicating superior positive predictive accuracy compared to other models and meeting the target performance criteria. Designed as a clinical tool, this model assists in selecting early embryo transfer or freezing on embryonic day 3 instead of blastocyst culture. Adjusting the decision threshold further personalizes the clinical application, enabling a focus on achieving a 90% positive predictive value (PPV) at 60% sensitivity or an 80% PPV at 80% sensitivity (Figure 3D).

### Subgroup Analyses

Subgroup analyses by facility for all models indicated no large differences between facilities, and the blastocyst formation prediction model was able to predict with an accuracy of AUC ≥0.83 for each facility (Figure 4A and Supplementary Figure 2A). Including age at oocyte retrieval as a feature, subgroup analyses by age group showed small differences between groups, and predictions were possible irrespective of the age group (Figure 4B and Supplementary Figure 2B). These findings highlight the robustness of our models.

**Figure 4.**
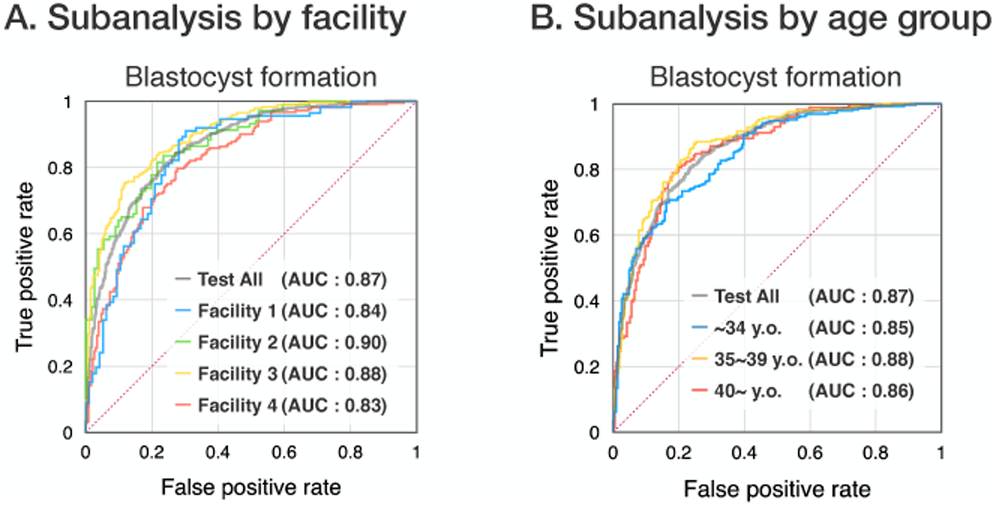
ROC subgroup analyses stratified by facility (**A**) and age group (**B**). AUC = area under the curve, ROC = receiver operating characteristic, y.o. = years old

### Comparison of Time-Lapse and Limited-Image Models

The AUC of the blastocyst formation prediction model built using XGBoost based on AI auto-annotated labels from single or three images and age was 0.63–0.78; however, the AUC of the model based on time-lapse image analysis was significantly higher, at 0.87 (p<1.0×10^-5^ for all comparisons; Figure 5A). The blastocyst formation prediction models based on image-only analysis by NASNet-A Large had an AUC of 0.64–0.76, and the prediction model based on time-lapse image analysis had a significantly higher AUC, at 0.87 (p<1.0×10^-5^ for all comparisons) (Figure 5B). The models based on time-lapse image analysis were also significantly better (p<1.0×10^-5^ for all comparisons) for good blastocyst formation prediction and poor blastocyst + arrested embryo prediction (Supplementary Figure 3).

**Figure 5.**
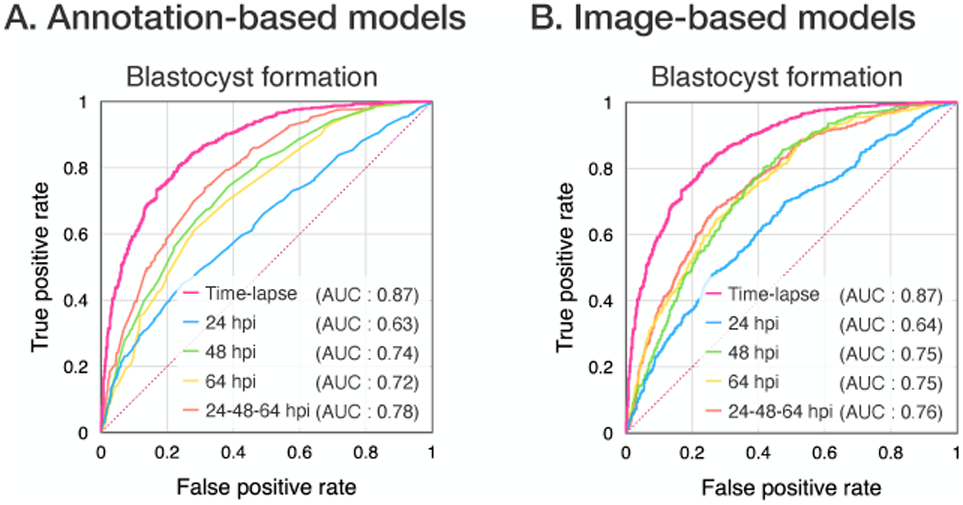
Blastocyst prediction models from single or few time-lapse images. (**A**) Comparison of time-lapse-based model with one or three label-based models. (**B**) Comparison of time-lapse-based model with one of three image-based models. AUC = area under the curve, hpi = hours post-insemination

### Feature Contribution and SHAP Analysis

The top 30 features for each model, ranked by absolute mean SHAP values (Figure 6A), consistently included proportions (2-cell stage, 4-cell stage, Veeck grade 2, etc.) and age at oocyte retrieval. While the time points of high contribution varied across models, a wide range of features within the 24–64 hpi broadly contributed to the models.

**Figure 6.**
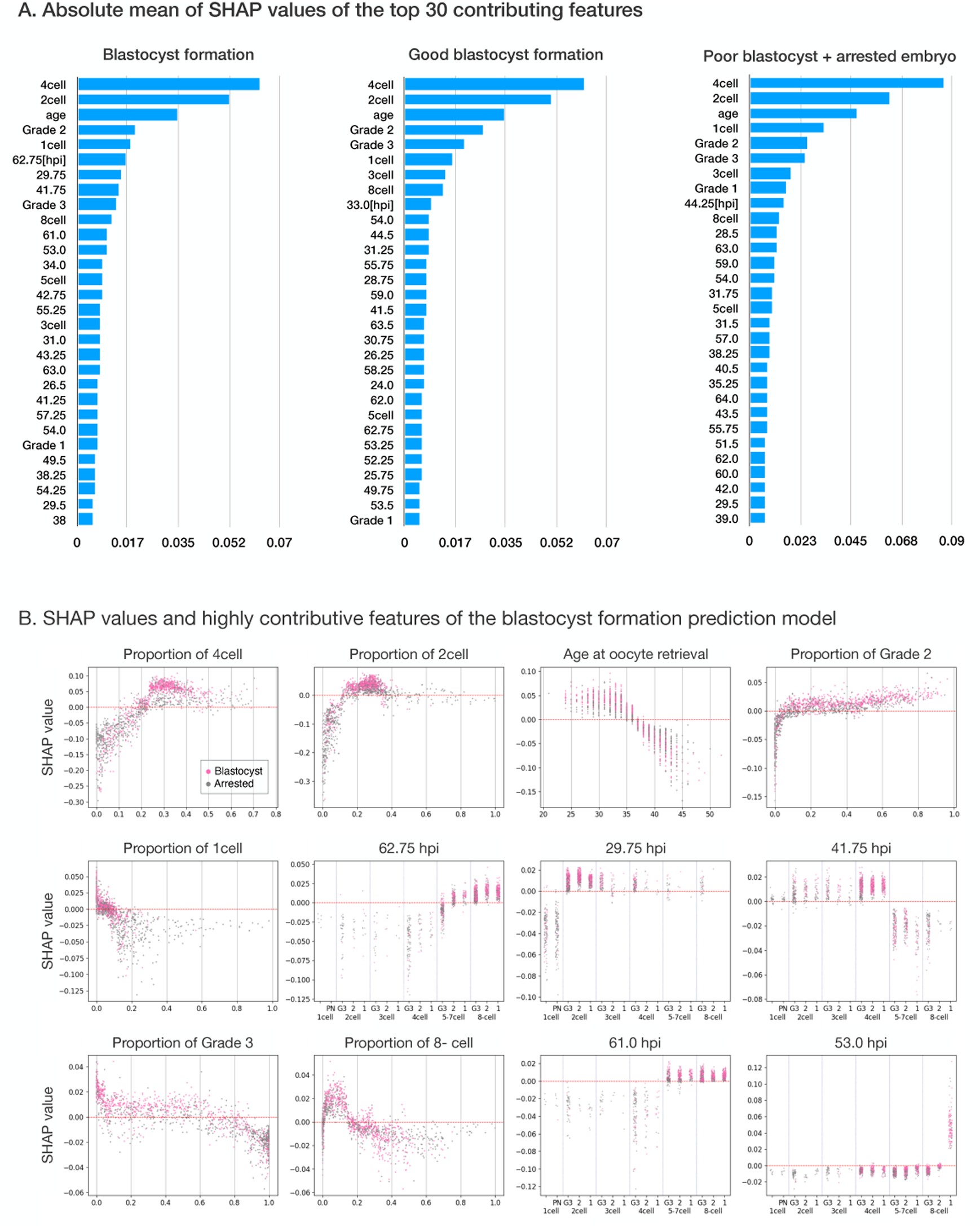
Visualization of model decision basis. (**A**) Top 30 contributing features based on SHAP. (**B**) SHAP values of highly contributing features of the blastocyst formation prediction model. SHAP values of highly contributing features of the good blastocyst formation and poor blastocyst + arrested embryo prediction models are shown in Supplementary Figure 4. SHAP = SHapley Additive exPlanations

The correlation between annotation labels and SHAP values (Figure 6B) at each time point in the blastocyst formation prediction model indicated that whether each feature contributes to a positive or negative prediction often varies at a single point because XGBoost deals with the 17-class labels as discrete-value variables rather than categorical variables. The 5–7-cell stage of grade 1/2 or 8-cell stage at 62.75 hpi, 2-cell stage or more at 29.75 hpi, and 4-cell stage or less at 41.75 hpi were found to contribute to positive predictions. Focusing on 41.75 hpi, the 5-cell or more stage due to overestimation of poor-grade embryos contributed to negative predictions, whereas the 3-cell or less stage, which was the most frequently arrested stage, incorrectly contributed to positive predictions. In contrast, at 42.75 hpi, the 3-cell or less stage contributed appropriately to negative predictions, and the entire model worked well.

The strong contribution of a very small proportion of 2-cell stage to negative predictions is thought to represent abnormal direct cleavage.

Although our AI auto-annotator occasionally overestimated the cell stage in poor-grade embryos, embryos with abnormally high proportion of 8-cell had negative SHAP values. Thus, these blastocyst prediction models were able to accurately evaluate even poor-grade embryos with morphological misclassification.

## Discussion

### Main Findings

In this study, 24–64-hpi time-lapse images recorded at various incubators and ages at oocyte retrieval from multiple centers were analyzed using AI to build models for predicting blastocyst morphology after the embryonic day 5. The AUCs for the blastocyst formation, good blastocyst formation, and poor blastocyst + arrested embryo prediction models were 0.87, 0.88, and 0.87, respectively. The positive predictive accuracy of the poor blastocyst + arrested embryo prediction model, which is envisioned as a clinical tool for deciding whether blastocyst culture should be continued, or early embryo freezing/transfer should be performed, achieved a PR AUC of 0.90, meeting the target accuracy.

### Performance and Versatility of the AI Auto-Annotator

The newly developed AI auto-annotator offers several advantages over previously published models. In particular, it is compatible with various types of incubators, attains the highest level of cell stage classification accuracy without labor-intensive manual annotation, and incorporates additional qualitative evaluations, including pronuclearity and Veeck grade. Our cell stage accuracy of 95% compares favorably with previous studies [40–44]. This performance was particularly remarkable considering that we included fragmented embryos (Veeck grade 3) and more difficult-to-classify stages such as 3- and 5-7-cell. A previous study reported a slightly higher accuracy [41], but their model included fewer classification categories and a limited set of embryos, which may not reflect real-world diversity. In contrast, our approach includes a broader range of embryo states, including those with fragmentation, making it more generalizable to clinical practice.

### Accuracy and Generalizability of the Prediction Models

We implemented versatile image pre-processing and classified time-lapse images at each time point using the highly accurate AI auto-annotator, enabling detailed analysis of stage and grade, calculation of parameters into robust prediction models.

Whereas prior day 3 blastocyst formation prediction models achieved validated ROC AUCs ranging from 0.73 to 0.82 [9,17,21,29], our model demonstrated superior performance with an AUC of 0.87. Similarly, our good blastocyst formation model achieved an ROC AUC of 0.88, improving on reported unvalidated AUCs of 0.73–0.82 [22–24]. Moreover, we successfully created a poor blastocyst + arrested embryo model with a PR AUC of 0.90, indicating its utility for identifying embryos unlikely to develop further. This prediction can guide clinicians in deciding whether to proceed with day 3 transfer or freezing the embryo rather than risk further culture.

Most previous models relied on time-lapse parameters or images captured within 24 hpi [11,12,22–24,29,13–18,20,21]. However, our study included only images of the embryos from 24 to 64 hpi, which are routinely recorded at most facilities, ensuring broader applicability while maintaining superior accuracy.

In contrast to previous AI models, which were device-dependent [25–27,29], our study analyzed images from four types of incubators across four facilities. By using a highly adaptable pre-processing method and a versatile AI auto-annotator, our models effectively managed variations such as aspect ratios, embryo area percentages, imaging contrasts, resolutions, and well geometries.

Data were collected irrespective of oocyte retrieval age, insemination method, or culture medium, creating heterogeneity across cases. Nonetheless, subgroup analyses showed minimal differences in AUC across facilities and age groups, confirming the model’s generalizability. These findings suggest that our models are versatile, robust, and suitable for diverse clinical settings.

### Interpretability of the prediction models

XGBoost outperformed Long Short-Term Memory models (data not shown) by analyzing features in parallel, effectively capturing temporal aspects of morphokinetic development from 24–64 hpi. SHAP analysis revealed how features, including human-recognizable parameters and embryo age, contributed to predictions. Importantly, localized misclassifications, such as overestimated cell stages in fragmented embryos, did not compromise overall model accuracy due to integration with more reliable features.

By restricting features to interpretable parameters, our approach ensures transparency and addresses the “black box” limitation often associated with AI. SHAP values allow clinicians to visualize individual embryo predictions, providing a clear understanding of the rationale behind the model’s decisions. This interpretability fosters clinician confidence, accelerates regulatory approval, and supports the clinical application of AI tools in embryo assessment.

### Limitations

Some limitations should be considered. First, we relied on the Gardner criteria [45,46] for blastocyst grading. While widely used, these criteria are subjective and prone to inter-rater variability [47]. To minimize domain shift, grading was conducted independently at each facility without standardization.

Second, embryos with missing records, misplacement, or poor-quality images were excluded, limiting absolute generalizability. However, this approach ensured a focus on clinically relevant embryos, enhancing practical applicability.

Finally, clinical heterogeneity in insemination methods, culture media, and patient demographics may have introduced variability. While our multicenter design mitigated these effects to some extent, further validation in prospective cohorts is warranted.

Despite these limitations, the study demonstrates the potential of AI models to advance embryo assessment in diverse clinical settings.

## Conclusions

In this study, we developed an interpretable AI model capable of accurately predicting blastocyst morphology based on time-lapse images taken between 24 and 64 hpi and maternal age. The model’s compatibility with various types of incubators ensures its broad applicability across diverse clinical settings. Notably, the poor blastocyst + arrested embryo prediction model offers significant clinical value, providing actionable insights for determining whether to continue blastocyst culture or perform early embryo freezing or transfer on day 3. By enabling embryo-specific decisions, this tool has the potential to optimize in vitro fertilization workflows, enhance individualized care, and improve patient outcomes.

## Data availability

The AI code and required weights for this study are available at GitHub (https://github.com/KU-ObGy-ART/blastocyst-prediction/). Owing to the nature of this research, the participants of this study did not agree that their data, including time-lapse images, should be shared publicly. Thus, images and clinical features of the fine-tuning and blastocyst prediction datasets are not available.

## Acknowledgments

The authors thank Dr. Aisaku Fukuda at the IVF Osaka Clinic, Dr. Kimihiko Nakamura at the Amanogawa Ladies Clinic, and Dr. Masahide Shiotani and Noritoshi Enatsu at the Hanabusa Women’s Clinic for generously providing the time-lapse images and clinical data.

## Author’s roles

**A Yanai:** Conceptualization, Methodology, Software, Investigation, Resources, Writing - Original Draft, Visualization, Formal analysis, Data Curation, Writing - Review & Editing. **A Horie:** Conceptualization, Methodology, Writing - Review & Editing, Project administration. **A Sakurai:** Investigation, Writing - Review & Editing. **S Imakita:** Investigation, Writing - Review & Editing. **M Nakamura:** Investigation, Writing - Review & Editing. **A Ikeda:** Investigation, Writing - Review & Editing. **S Shitanaka:** Validation, Writing - Review & Editing. **T Ohara:** Validation, Writing - Review & Editing. **B Nakakita:** Conceptualization, Methodology, Writing - Review & Editing. **A Ueda:** Methodology, Software, Resources, Writing - Review & Editing. **Y Kitawaki:** Investigation, Writing - Review & Editing. **Y Sagae:** Methodology, Funding acquisition, Writing - Review & Editing. **A Okunomiya:** Methodology, Resources, Writing - Review & Editing, **M Mandai:** Supervision, Writing - Review & Editing.

---

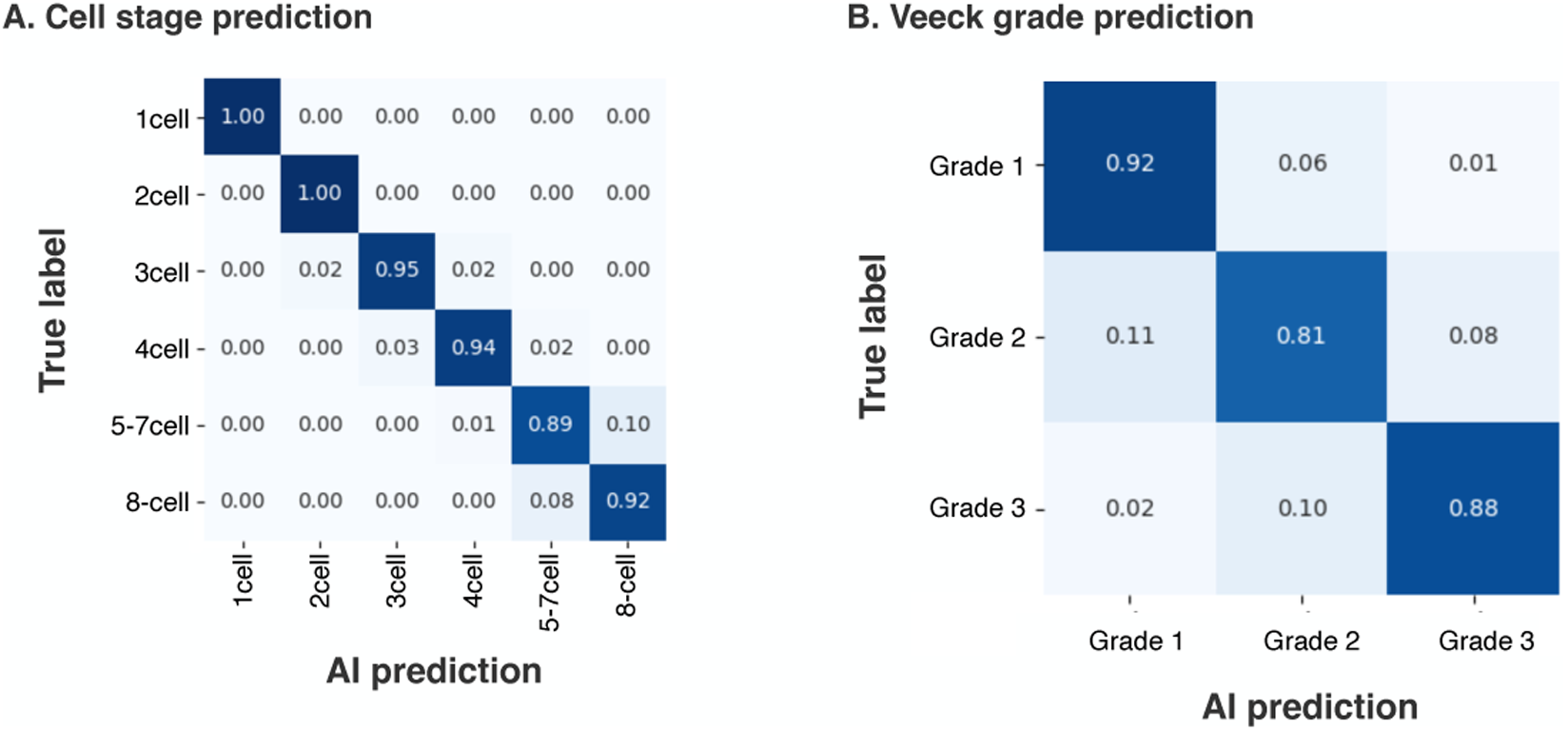

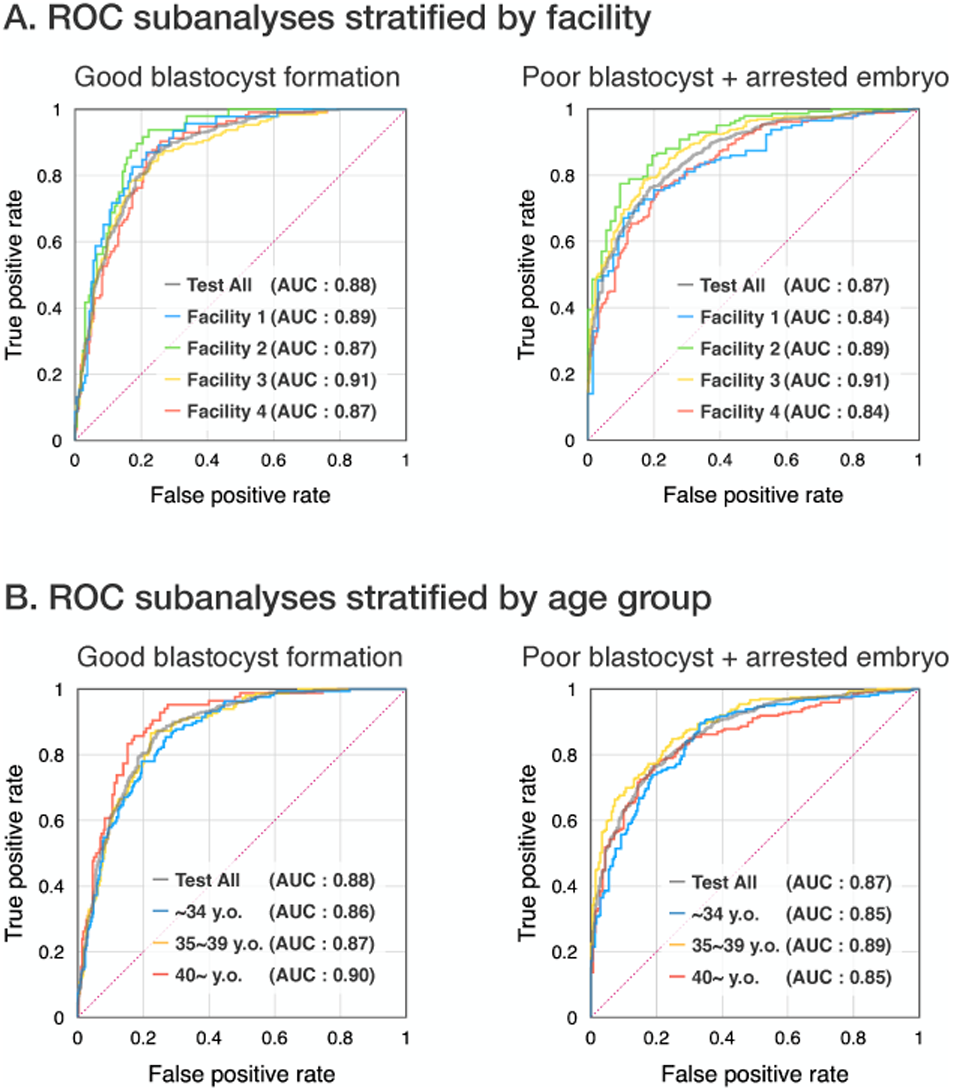

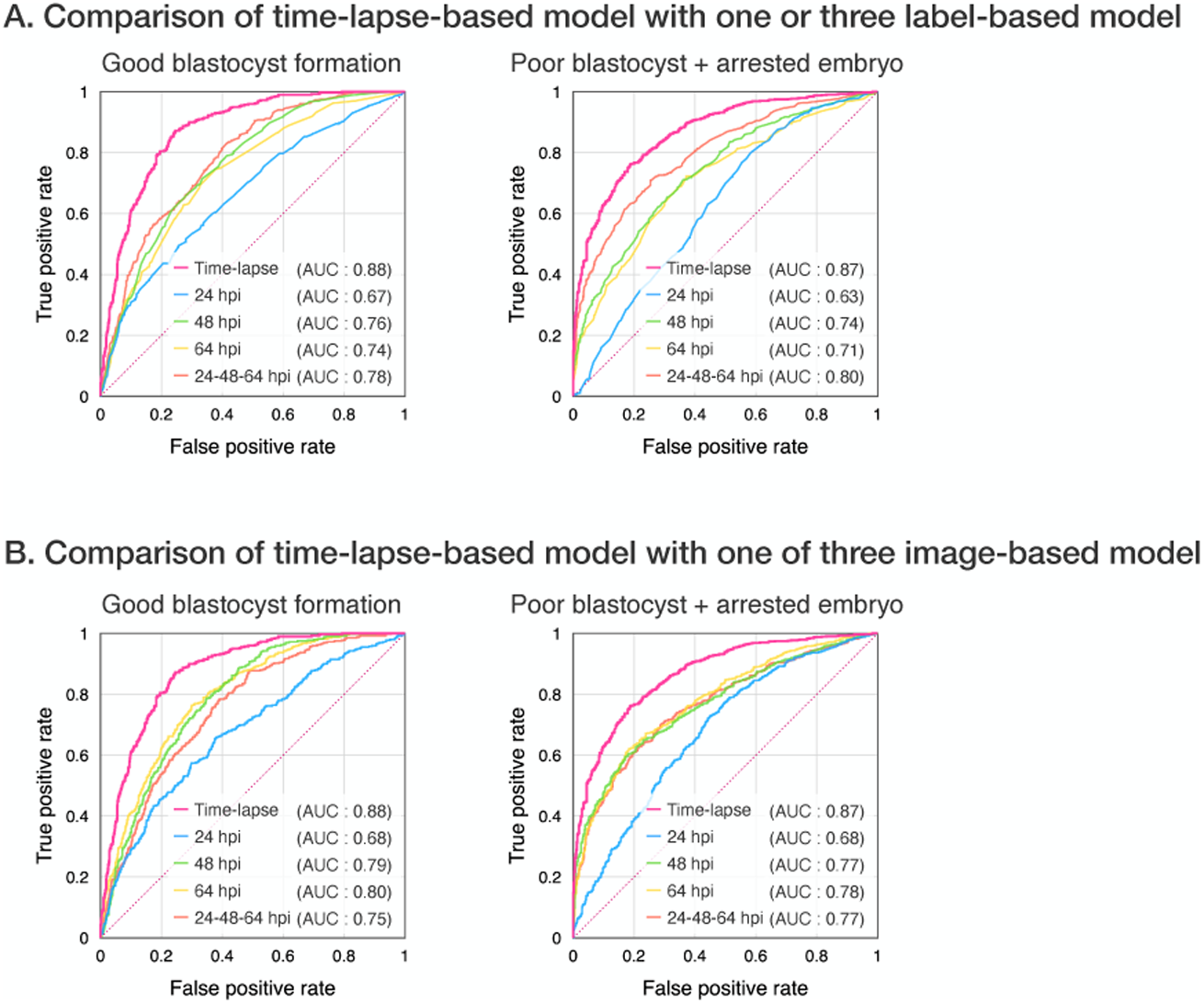

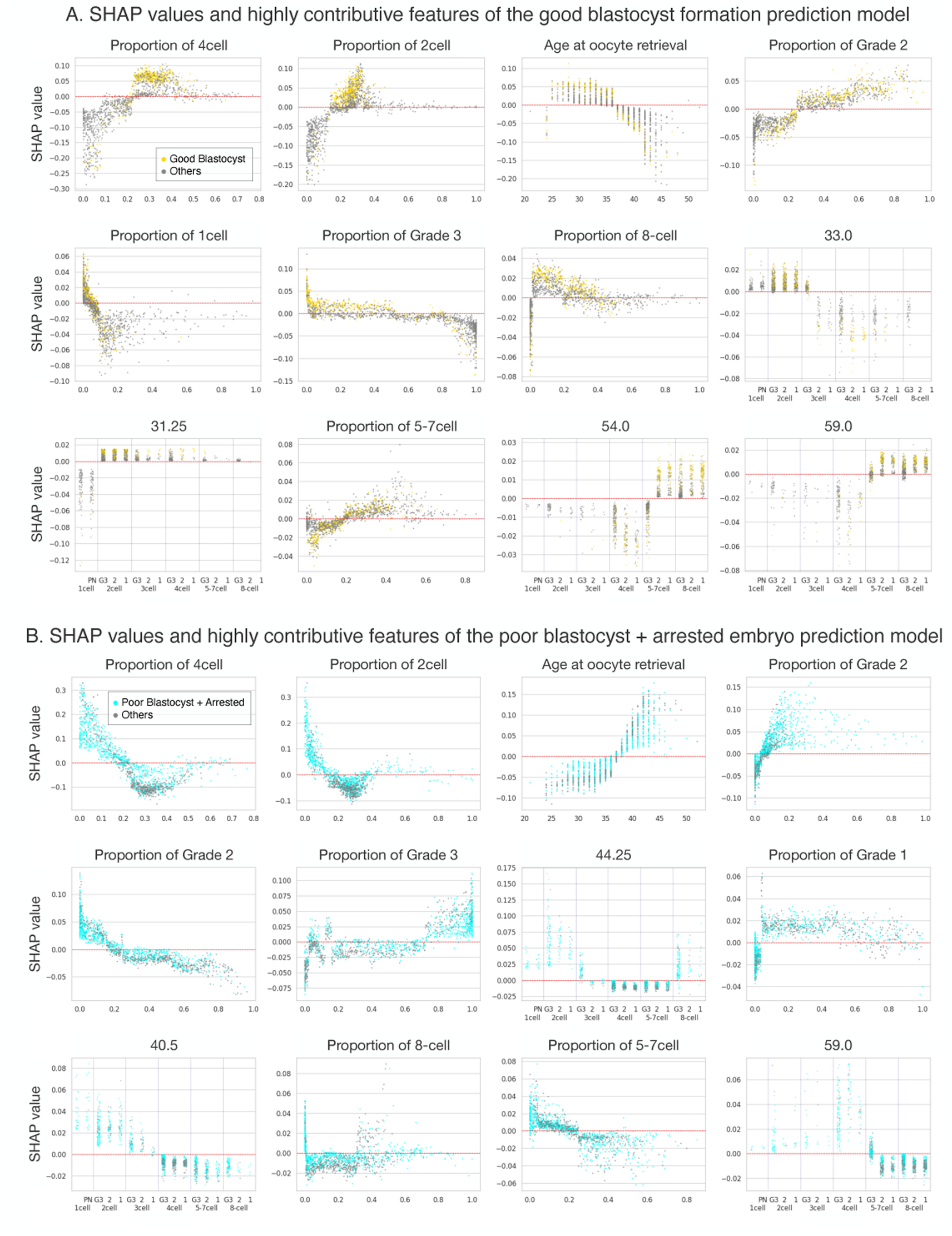

