## Supplementary Text 1 for "Innovative AI models for clinical decision-making: predicting blastocyst formation and quality from time-lapse embryo images up to embryonic day 3"

**Artificial intelligence (AI) method**

**Machine and environment**

Ubuntu 18.04.6 LTS, Xeon (Broadwell) 2.2 GHz 4core, 12.6 GB memory, GPU (Tesla V100 16GB, NVIDIA) on Google Colaboratory, Python 3.7.15, OpenCV 4.6.0.66, TensorFlow 2.9.2, Scikit-learn 1.0.2.

**Fine-tuning of the AI auto-annotator**

Morphologically classified time-lapse and stereomicroscopic images were preprocessed using the Python script (https://github.com/KU-ObGy-ART/blastocyst-prediction/preprocess.ipynb) using the OpenCV library to create supervised images.

During preprocessing, the image levels were adjusted, outline of the embryo area fitted with a circumscribed ellipse, and area cropped into a square whose sides were the averages of the short and long diameters. The cropped image was resized to 331 × 331 pixels and circularly masked with black around the perimeter.

More than 1,000 supervised images from more than 1,000 embryos in each group of 17 classes were preprocessed (Figures 1B and 2A) and split train: validation: test = 8:1:1 to create a fine-tuning dataset. The ImageNet-1k pre-trained NASNet-A Large implemented with TensorFlow 2 was trained using this dataset. The training images were randomly flipped, rotated, zoomed, and shifted by online data augmentation using ImageDataGenerator(horizontal_flip=True, rotation_range=180, zoom_range=0.1, width_shift_range=30, height_shift_range=30, fill_mode='constant'). The loss function was a cross-entropy loss, the maximum epoch was 100, the optimizer was Adam, the learning rate was started at 1E-5 and decreased by 1/10 for every four epochs, the loss function did not decrease, and the early stopping period was 16 epochs.

**Data collection for the blastocyst prediction dataset**

The data collection periods differed by facility, with Facility 1 collecting data from May 2018 to May 2022; Facility 2, from March 2019 to November 2021; Facility 3, from April 2020 to June 2022; and Facility 4, from January 2021 to December 2022. Each facility had a different time-lapse incubator, with facility 1 using CCM-iBIS (ASTEC, Kyoto, Japan), facility 2 using Geri (Genea Biomedix, Sydney, Australia), facility 3 using EmbryoScope+ (Vitrolife, Göteborg, Sweden), and facility 4 using CCM-iBIS-SG (ASTEC, Kyoto, Japan).

Time-lapse cultures were started from embryonic day 1 for conventional in vitro fertilization and from day 0 for ICSI at Facility 1, day 1 at Facility 2, and day 0 at Facilities 3 and 4. Time-lapse images were recorded every 5 min at Facility 2, every 10 min at Facility 3, and every 15 min at Facilities 1 and 4.

Time-lapse images were collected every 15 min between 24–64 hours post-insemination (hpi), with an error margin of 8 min.

**Preprocessing for the blastocyst prediction dataset**

Images of 7-11 focal axes were recorded by the incubator at each time point, and images of the central axis were used. Time-lapse images were recorded as JPG images or mp4 movies, and the movies were converted to JPG images for each frame. Time-lapse images were preprocessed using a Python scrip (https://github.com/KU-ObGy-ART/blastocyst-prediction/preprocess.ipynb) using the OpenCV library and images from the fine-tuned dataset. The embryo area was cropped from the image, resized to 331 × 331 pixels, and circularly masked black.

Embryos were excluded if more than 5% of the image data was missing per sample due to missing records, large debris with a long diameter exceeding 5% of the image size, embryo misplacement, or errors in image preprocessing. Additionally, poor-quality embryos classified as Veeck grade 4 or 5 and degenerated embryos were also excluded.

**Blastocyst prediction AI build**

Time-lapse images at each time point were morphologically classified using an artificial intelligence auto-annotator. Classification labels, the proportion of each cellular stage, Veeck grade, and age at egg retrieval were used as features. The model was built by analyzing the dataset for training and cross-validation using scikit-learn and XGBoost.

XGBoost hyperparameters were searched for optimal values by GridSearchCV (cv=5, scoring='balanced_accuracy') in scikit-learn. The search ranges were learning_rate [0.01, 0.05, 0.10], max_depth [4, 5, 6, 7, 8, 9, 10], and n_estimators [50, 60, 80, 100, 120, 200]. We set rondom_state to 0 and used the default values for the other hyperparameters.

The number of features was reduced and optimized using recursive feature elimination.

**Supplementary Figure 1.** Confusion matrix of AI auto-annotator prediction. (**A**) Cell stage prediction. (**B**) Veeck grade prediction.

**Supplementary Figure 2.** ROC subanalysis. (**A**) ROC subanalysis by facility. (**B**) ROC subanalysis by age group. ROC = receiver operating characteristic

**Supplementary Figure 3.** Comparison of prediction models from time-lapse images with single or few images. (**A**) Comparison of the time-lapse-based model with one or three label-based models. (**B**) Comparison of the time-lapse model with one of three image-based models. ROC = receiver operating characteristic

**Supplementary Figure 4**. SHAP values of highly contributing features. (**A**) Good blastocyst formation prediction model. (**B**) Poor blastocyst + arrested embryo prediction model. SHAP = SHapley Additive exPlanations

**Supplementary Table 1.** Comparison between AI-calculated and previously reported time-lapse parameters. Previous reports [1–5] show the mean or median ranges. Median (quartile range), *Mann–Whitney U test

**Supplementary Table 2.** Feature list and XGBoost hyperparameters of prediction models. (**A**) Feature list of the blastocyst formation prediction model. (**B**) Feature list of the good blastocyst prediction model. (**C**) Feature list of the poor blastocyst + arrest prediction models. (**D**) XGBoost hyperparameters of the prediction models
