## Supplementary Table 1 for "Innovative AI models for clinical decision-making: predicting blastocyst formation and quality from time-lapse embryo images up to embryonic day 3"

**Supplementary Table 1 Comparison of AI-calculated time-lapse parameters with those previously reported**

|  | All (n = 7,111) | Blastocyst (n = 3,579) | Previous reports | Arrested (n = 3,532) | Previous reports | *p* value (Blastocyst vs. Arrested) |
| --- | --- | --- | --- | --- | --- | --- |
| t2 [hpi] | 25.8 [24.0–28.5] | 24.8 [24.0–26.8] | 23.7– 27.6 | 27.0 [24.5–30.3] | 24.7–30.1 | <0.001 |
| t3 [hpi] | 33.0 [28.3–37.3] | 33.3 [28.3–36.8] | 33.1–39.2 | 33.0 [28.5–38.5] | 33.1–40.8 | <0.001 |
| t4 [hpi] | 34.8 [30–38.8] | 35.3 [31.5–38.3] | 36.0–39.9 | 34.0 [29.0–40.0] | 37.6–42.4 | 0.01 |
| t3-t2 [hr] | 7.5 [1.5–11.3] | 9.0 [2.0–11.3] | 9.4–12.4 | 4.0 [1.3–11.3] | 9.8–13.0 | <0.001 |
| t4-t3 [hr] | 0.8 [0.3–2.3] | 0.8 [0.3–2.5] | 0.6–1.8 | 0.5 [0.0–2.3] | 0.8–3.3 | <0.001 |

hpi: hours post-insemination
