## Supplementary Table 2 for "Innovative AI models for clinical decision-making: predicting blastocyst formation and quality from time-lapse embryo images up to embryonic day 3"

**Supplementary Table 2 Features list and XGBoost hyper-parameter of prediction models**

A. Features list of blastocyst formation prediction model

| 24.0 hpi | 32.25 hpi | 42.75 hpi | 56.25 hpi | Age at oocyte retrieval |
| --- | --- | --- | --- | --- |
| 25.0 hpi | 32.75 hpi | 43.25 hpi | 56.5 hpi | Proportion of 1cell |
| 25.25 hpi | 33.0 hpi | 43.5 hpi | 57.0 hpi | Proportion of 2cell |
| 25.5 hpi | 33.25 hpi | 43.75 hpi | 57.25 hpi | Proportion of 3cell |
| 26.0 hpi | 34.0 hpi | 44.0 hpi | 57.5 hpi | Proportion of 4cell |
| 26.5 hpi | 34.75 hpi | 44.25 hpi | 57.75 hpi | Proportion of 5-7cell |
| 26.75 hpi | 35.0 hpi | 44.5 hpi | 58.75 hpi | Proportion of 8-cell |
| 27.5 hpi | 35.5 hpi | 44.75 hpi | 59.25 hpi | Proportion of grade 1 |
| 27.75 hpi | 37.0 hpi | 45.5 hpi | 60.0 hpi | Proportion of grade 2 |
| 28.25 hpi | 38.0 hpi | 47.0 hpi | 60.25 hpi | Proportion of grade 3 |
| 28.5 hpi | 38.25 hpi | 47.25 hpi | 60.5 hpi |  |
| 29.25 hpi | 38.75 hpi | 47.75 hpi | 61.0 hpi |  |
| 29.5 hpi | 39.25 hpi | 48.0 hpi | 61.75 hpi |  |
| 29.75 hpi | 39.5 hpi | 49.5 hpi | 62.0 hpi |  |
| 30.5 hpi | 40.25 hpi | 49.75 hpi | 62.75 hpi |  |
| 30.75 hpi | 40.5 hpi | 50.5 hpi | 63.0 hpi |  |
| 31.0 hpi | 41.25 hpi | 53.0 hpi | 63.5 hpi |  |
| 31.25 hpi | 41.5 hpi | 54.0 hpi | 64.0 hpi |  |
| 31.5 hpi | 41.75 hpi | 54.25 hpi |  |  |
| 32.0 hpi | 42.25 hpi | 55.25 hpi |  |  |

B. Features list of good blastocyst prediction model

| 24.0 hpi | 31.75 hpi | 41.0 hpi | 49.25 hpi | 57.5 hpi | Age at oocyte retrieval |
| --- | --- | --- | --- | --- | --- |
| 24.25 hpi | 32.0 hpi | 41.25 hpi | 49.5 hpi | 57.75 hpi | Proportion of 1cell |
| 24.5 hpi | 32.25 hpi | 41.5 hpi | 49.75 hpi | 58.0 hpi | Proportion of 2cell |
| 24.75 hpi | 32.5 hpi | 41.75 hpi | 50.0 hpi | 58.25 hpi | Proportion of 3cell |
| 25.0 hpi | 32.75 hpi | 42.0 hpi | 50.25 hpi | 58.5 hpi | Proportion of 4cell |
| 25.25 hpi | 33.0 hpi | 42.25 hpi | 50.5 hpi | 58.75 hpi | Proportion of 5-7cell |
| 25.5 hpi | 33.25 hpi | 42.5 hpi | 50.75 hpi | 59.0 hpi | Proportion of 8-cell |
| 25.75 hpi | 33.5 hpi | 42.75 hpi | 51.0 hpi | 59.25 hpi | Proportion of grade 1 |
| 26.0 hpi | 33.75 hpi | 43.0 hpi | 51.25 hpi | 59.75 hpi | Proportion of grade 2 |
| 26.25 hpi | 34.25 hpi | 43.25 hpi | 51.5 hpi | 60.0 hpi | Proportion of grade 3 |
| 26.5 hpi | 34.75 hpi | 43.5 hpi | 51.75 hpi | 60.25 hpi |  |
| 26.75 hpi | 35.0 hpi | 43.75 hpi | 52.0 hpi | 60.5 hpi |  |
| 27.0 hpi | 35.5 hpi | 44.0 hpi | 52.25 hpi | 60.75 hpi |  |
| 27.25 hpi | 35.75 hpi | 44.25 hpi | 52.5 hpi | 61.0 hpi |  |
| 27.5 hpi | 36.0 hpi | 44.5 hpi | 52.75 hpi | 61.25 hpi |  |
| 27.75 hpi | 36.25 hpi | 44.75 hpi | 53.0 hpi | 61.5 hpi |  |
| 28.0 hpi | 36.5 hpi | 45.0 hpi | 53.25 hpi | 61.75 hpi |  |
| 28.25 hpi | 36.75 hpi | 45.25 hpi | 53.5 hpi | 62.0 hpi |  |
| 28.5 hpi | 37.0 hpi | 45.5 hpi | 53.75 hpi | 62.25 hpi |  |
| 28.75 hpi | 37.25 hpi | 45.75 hpi | 54.0 hpi | 62.5 hpi |  |
| 29.0 hpi | 37.5 hpi | 46.0 hpi | 54.25 hpi | 62.75 hpi |  |
| 29.25 hpi | 38.25 hpi | 46.25 hpi | 54.5 hpi | 63.0 hpi |  |
| 29.5 hpi | 38.5 hpi | 46.5 hpi | 54.75 hpi | 63.25 hpi |  |
| 30.0 hpi | 38.75 hpi | 46.75 hpi | 55.0 hpi | 63.5 hpi |  |
| 30.25 hpi | 39.0 hpi | 47.25 hpi | 55.25 hpi | 63.75 hpi |  |
| 30.5 hpi | 39.5 hpi | 47.5 hpi | 55.5 hpi | 64.0 hpi |  |
| 30.75 hpi | 39.75 hpi | 47.75 hpi | 55.75 hpi |  |  |
| 31.0 hpi | 40.25 hpi | 48.5 hpi | 56.5 hpi |  |  |
| 31.25 hpi | 40.5 hpi | 48.75 hpi | 57.0 hpi |  |  |
| 31.5 hpi | 40.75 hpi | 49.0 hpi | 57.25 hpi |  |  |

C. Features list of poor blastocyst + arrested embryo prediction model

| 24.25 hpi | 35.5 hpi | 50.75 hpi | 61.0 hpi | Age at oocyte retrieval |
| --- | --- | --- | --- | --- |
| 24.75 hpi | 38.25 hpi | 51.5 hpi | 62.0 hpi | Proportion of 1cell |
| 26.5 hpi | 39.0 hpi | 52.0 hpi | 62.75 hpi | Proportion of 2cell |
| 27.5 hpi | 39.5 hpi | 52.5 hpi | 63.0 hpi | Proportion of 3cell |
| 27.75 hpi | 40.5 hpi | 53.0 hpi | 63.5 hpi | Proportion of 4cell |
| 28.5 hpi | 41.25 hpi | 53.25 hpi | 64.0 hpi | Proportion of 5-7cell |
| 29.25 hpi | 41.75 hpi | 54.0 hpi |  | Proportion of 8-cell |
| 29.5 hpi | 42.0 hpi | 54.25 hpi |  | Proportion of grade 1 |
| 29.75 hpi | 42.25 hpi | 54.5 hpi |  | Proportion of grade 2 |
| 30.5 hpi | 42.75 hpi | 54.75 hpi |  | Proportion of grade 3 |
| 30.75 hpi | 43.25 hpi | 55.25 hpi |  |  |
| 31.25 hpi | 43.5 hpi | 55.75 hpi |  |  |
| 31.5 hpi | 43.75 hpi | 56.25 hpi |  |  |
| 31.75 hpi | 44.25 hpi | 56.5 hpi |  |  |
| 32.0 hpi | 48.5 hpi | 57.0 hpi |  |  |
| 32.5 hpi | 48.75 hpi | 58.25 hpi |  |  |
| 33.0 hpi | 49.25 hpi | 59.0 hpi |  |  |
| 33.25 hpi | 49.5 hpi | 59.75 hpi |  |  |
| 34.5 hpi | 49.75 hpi | 60.0 hpi |  |  |
| 35.25 hpi | 50.5 hpi | 60.25 hpi |  |  |

D. XGBoost hyper-parameter of prediction models

| **Model** | learning_rate | max_depth | n_estimators |
| --- | --- | --- | --- |
| Blastocyst formation prediction | 0.05 | 8 | 100 |
| Good blastocyst prediction | 0.05 | 5 | 200 |
| Poor blastocyst + arrested embryo prediction | 0.05 | 5 | 200 |

hpi: hours post-insemination, grade: Veeck’s grade
